# Underestimation of SARS-CoV-2 in wastewater due to single or double mutations in the N1 qPCR probe binding region

**DOI:** 10.1101/2024.02.03.24302274

**Authors:** Jianxian Sun, Minqing Ivy Yang, Jiaxi Peng, Ismail Khan, Jhoselyn Jaramillo Lopez, Ronny Chan, Elizabeth Edwards, Hui Peng

## Abstract

Wastewater surveillance using RT-qPCR has now been widely adopted to track circulating levels of SARS-CoV-2 virus in many sewer sheds. The CDC qPCR assays targeting two regions (N1 and N2) within the N gene are commonly used, but a discrepancy between the two biomarkers has been noticed by many groups using this method since late 2021. The reason is presumed to be due to mutations in regions targeted by the qPCR probe. In this study, we systematically investigated and unequivocally confirmed that the underlying reason for this discrepancy was mutations in the N1 probe target, and that a single mutation could cause a significant drop in signal. We first confirmed the proportion of related mutations in wastewater samples (Jan 2021-Dec 2022) using nested PCR and LC-MS. Based on relative proportion of N1 alleles, we separated the wastewater data into four time periods corresponding to different variant waves: Period I (Alpha and Delta waves with 0 mutation), Period II (BA.1/BA.2 wave with a single mutation found in all Omicron strains), Period III (BA. 5.2* wave with two mutations), and Period IV (BQ.1* wave with two mutations). Significantly lower N1 copies relative to N2 copies in samples from Periods II-IV compared to those from Period I was observed in wastewater. To further pinpoint the extent to which each mutation impacted N1 quantification, we compared the qPCR response among different synthetic oligomers with corresponding mutations. This study highlighted the impact of even just one or two mutations on qPCR-based wastewater surveillance of SARS-CoV-2.

## 1. Introduction

The outbreak of a novel coronavirus pneumonia (COVID-19) led to over 4.77 million positive cases and 54,498 deaths in Canada by October 24, 2023(Government-of-Canada 2023). Due to the high cost and limited capacity associated with nasopharyngeal swab testing, most countries including Canada discontinued population-wide clinical polymerase chain reaction (PCR) testing in 2022. Wastewater-based epidemiology (WBE) has now become the primary method to assess population-level trend of SARS-CoV-2 (Kirby et al. 2021, Peccia et al. 2020, Zheng et al. 2022). Compared to testing individuals using nasopharyngeal swab PCR, WBE proves to be a cost-effective population-level solution, offering early warning information ahead of clinical data (Galani et al. 2022, Peccia et al. 2020). Recent research has demonstrated a close correlation between the trends of SARS-CoV-2 abundance in wastewater and clinical data (D’Aoust et al. 2022, Nourbakhsh et al. 2022, Peccia et al. 2020, Pileggi et al. 2022), underscoring the effectiveness of wastewater surveillance in monitoring SARS-CoV-2.

Reverse transcription quantitative PCR (RT-qPCR) has been the workhorse for SARS-CoV-2 wastewater surveillance due to its low cost, high sensitivity, and accuracy. However, the virus has been mutating since its emergence in late 2019 as indicated in sequences shared on the publicly available platform GISAID (https://gisaid.org/). Some variants are classified as variants of concern (VOCs) by WHO based on their phenotype and impact on countermeasures. A major concern for qPCR-based wastewater surveillance is that if mutations occur in any region targeted by the assay, it might lead to inaccuracies in quantification. The US Centers for Disease Control (CDC) N1 and N2 assays targeting two subregions of the N gene encoding the nucleocapsid protein (Lu et al. 2020) are the most widely used targets, because they are less prone to mutations compared to other regions of the genome, such as the S gene (https://gisaid.org/lineage-comparison/). The CDC assay targets two distinct regions of the virus to provide additional confidence in the specificity and accuracy of the results. As both targets are to the same gene, they are expected to be present at a 1:1 ratio. Most of the early VOCs including Alpha, Beta, Gamma, and Delta variants have no mutations in the CDC N1 or N2 regions. However, Omicron and its sub-lineages which started circulating worldwide in late December 2021, had several mutations in the N1 probe binding region while had no mutation reported in primer regions (Peng et al. 2022). These mutations can be seen in an alignment based on sequences shared on GISAID (Fig. 1 and Table S2). Notably, the C to T point mutation at position 28311 (C28311T) of the N1 probe is present in all Omicron sub-linages. Moreover, additional mutations appeared in the N1 probe region in certain BA.5 sub-linages including the A28330G mutation in BA.5.2/BF.7 strains, and the C28312T mutation in BQ.1/BQ.1.1 strains (Fig. 1). The presence of these point mutations raised some concerns over the quantification accuracy of qPCR based WBE, although there is considerable debate as to the impact of such small sequence changes.

**Fig. 1.**
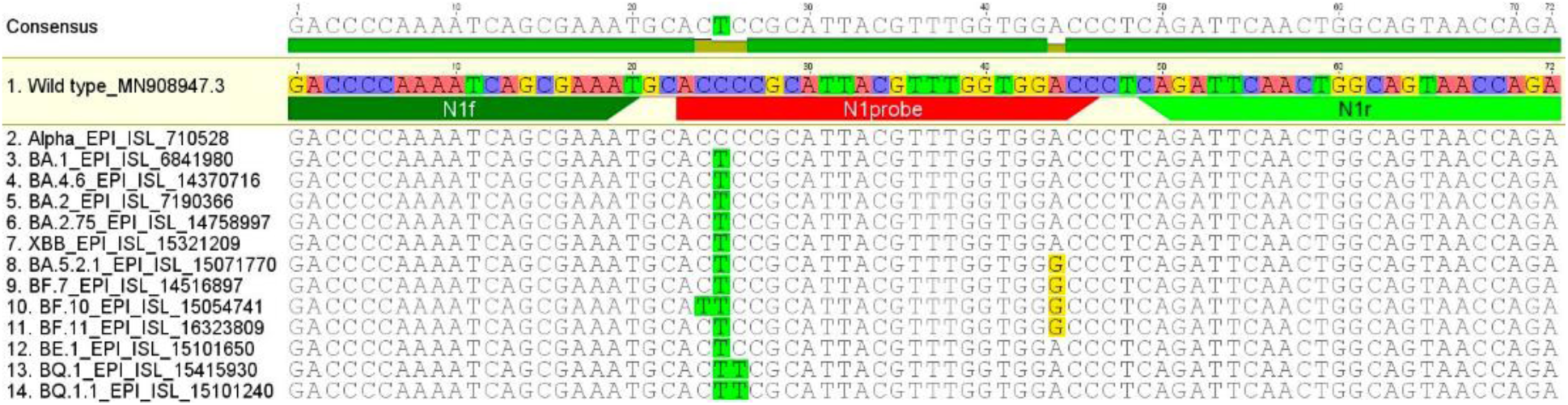
Alignment of CDC N1 assay region with sequences reported in Omicron variants.

In our routine SARS-CoV-2 WBE program at the University of Toronto, we noticed a shift in the ratio of N1 to N2 since the outbreak of Omicron in late 2021 in samples from all Toronto municipal wastewater treatment plants. This led us to systematically investigate the impact of mutations in the N1 probe binding region on RT-qPCR quantification. Using weekly wastewater samples from Toronto Ashbridges Bay (TAB, Toronto Ontario), we measured the abundance of mutated sequences using an orthologous and previously described liquid chromatography-mass spectrometry (LC-MS) method and compared this analysis with N1 and N2 quantification based on RT-qPCR. We also designed four synthetic oligomers representing specific variant strains and measured the magnitude of their impact in RT-qPCR reactions. These data unequivocally confirm that the observed “N1 drop-out” or discrepancy between N1 and N2 signals in RT-qPCR-based surveillance was due to strain-dependent mutations in the probe region.

## 2. Results and Discussion

### 2.1. Temporal trends of N1 and N2 concentrations in Toronto wastewaters

SARS-CoV-2 levels in the raw influents from TAB were monitored beginning Feb 11, 2021. As shown in Fig. 2A, both N1 (blue circles) and N2 (green diamonds) concentrations (copy/mL) in TAB wastewater fluctuated during the two years, mainly driven by the waves of different SARS-CoV-2 variants such as B.1.1.7 (Alpha), B.1.617.2 (Delta), and various B.1.1.529 (Omicron) sub-lineages that circulated in Canada (cov-spectrum.org). N1 and N2 are two biomarkers of SARS-CoV-2, so they are expected to be present at approximately 1:1 ratio in any sample with a reasonably intact N gene in the extracted RNA. For RT-qPCR absolute quantification, the stability of standards during storage and potential human errors in the calibration curve preparation are common issues that can adversely affect the accuracy of quantification. Since the same standard, which contained both N1 and N2 gene sequences on the same template string with 1:1 ratio (N-plasmid or ConcatP), was used for both N1 and N2 quantifications throughout the two years of surveillance, we expected a consistent N2/N1 ratio in wastewater samples. Consistent with our expectation, N2/N1 ratios in RT-qPCR were detected close to 1 in TAB wastewaters during Alpha and Delta waves (Feb 2021 to Dec 2021, Fig. 2C). However, we started to observe an increase in the N2/N1 ratio when Omicron became dominate after Dec 2021 (Fig. 2C). Since limited mutations in the N2 region were reported (Hasan et al. 2021) or sequences from GISAID, these results implicated that reported mutations in the N1 region of Omicron variants might be responsible for the underestimation of N1 gene copies in wastewaters in our RT-qPCR. The mutations in N1 probe region would potentially affect the binding affinity of the probe to the template, and thus affect N1 quantification in wastewater when using the wildtype sequence as standard in RT-qPCR.

**Fig. 2.**
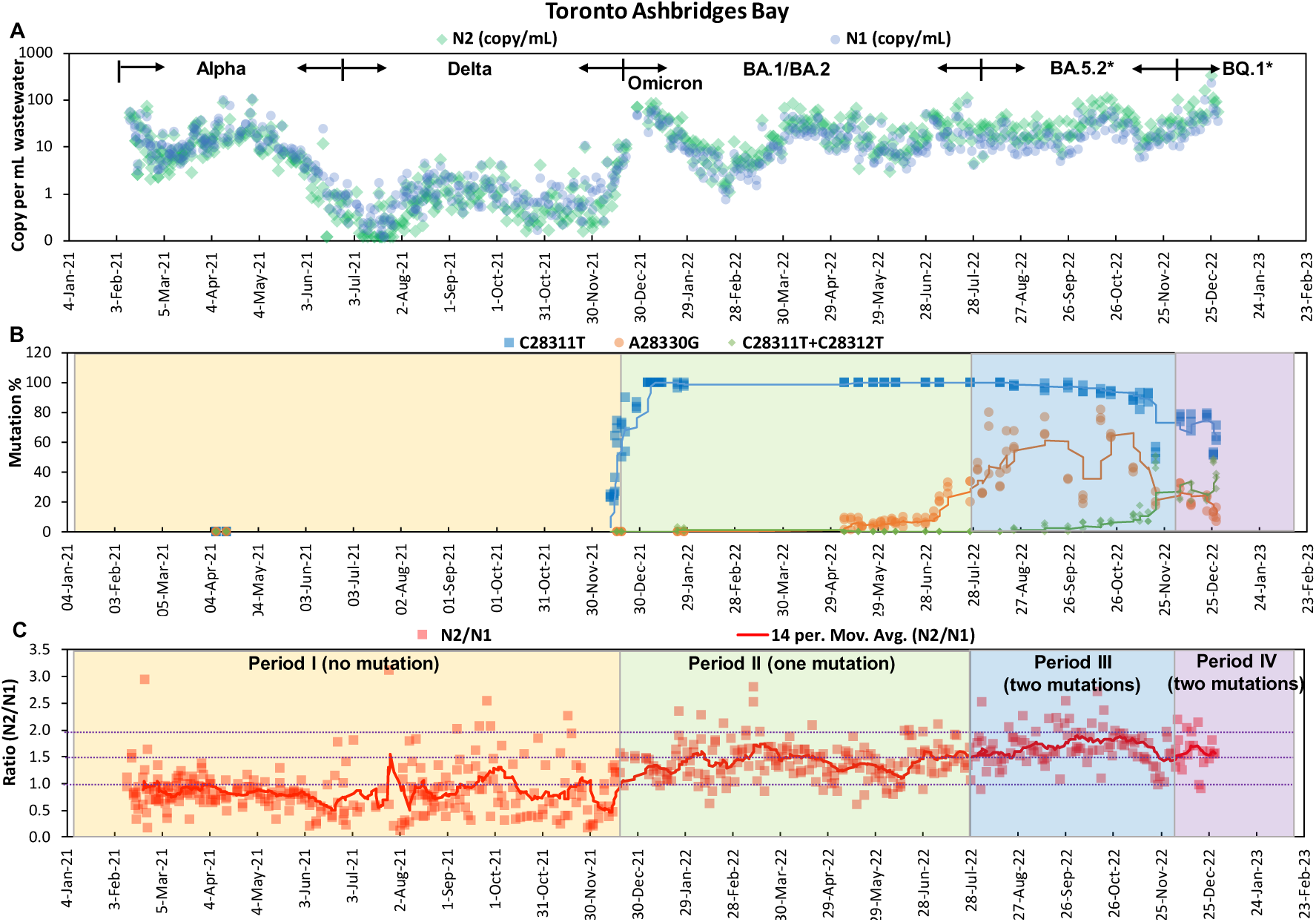
Trends of N1 (green dots) and N2 concentrations (blue dots) (copies/mL wastewater) (A), all different mutation patterns in N1 probe region (B), and N2/N1 ratios with 14-day moving average (C) in TAB wastewater. The proportions for C28311T in Fig. 2B during Dec 12, 2021 to Jan 27, 2022 were from our previous paper (Peng et al. 2022). Proportions of each mutation analyzed by LC-MS in Fig. 2B were nPCR products amplified from N1 in Fig. 2A. Background colors in Fig. 2B and 2C indicated different periods, which were separated based on mutation patterns in Fig. 2B.

### 2.2. Mass Spectrometry (nPCR-LC-MS) to confirm actual proportions of variant N1 sequences present in wastewater samples

To accurately quantify the proportions of different variant N1 sequences in wastewater samples, we employed a nested PCR-LC-MS method previously developed in our group (Peng et al. 2022). This LC-MS-based method can discriminate single-base mutations that are challenging to differentiate by allele-specific qPCR methods. By using two sets of nested PCR reactions (primer sequences shown in Table S3), mutations located at positions 28310-28312 and 28328-28330 within the N1 probe binding region were amplified and analyzed separately by LC-MS. Three genotypes within the 28310-28312 region could be detected in selected wastewater samples, which were CCC (wildtype), CTC (universal to all Omicron), and CTT (BQ.1*)/TTC (BF.10) (Fig. 2B). Two genotypes in the 28328-28330 region were detected which were GGA (wildtype) and GGG (BA.5.2* including BF.7 and BF.10) (Fig. 2B). Some mutations could not be distinguished by LC-MS, such as the CTT mutation (in BQ.1*) vs. TTC mutation (in BF.10) in the 28310-28312 amplicon, because these two mutations share the same *m/z* and have similar retention time in HPLC. However, BQ.1* variants with the CTT mutation circulated at a much higher rate than BF.10 with the TTC mutation (Public Health Ontario, 2022). Hence, it was safe to assume that the detected CTT+TTC mutation would predominantly represent BQ.1*.

To investigate the effects of those mutations on N1 quantification in wastewater, we separated the 2-year long wastewater surveillance data (Feb 11, 2021 to Dec 28, 2022) into four different periods according to the mutation patterns determined by nPCR-LC-MS (Fig. 2B). In Period I (Feb 11, 2021-Dec 16, 2021) corresponding to Alpha and Delta waves, no mutation in the N1 probe binding region was detected in wastewater samples. In Period II (Dec 17, 2021 - Jul 31, 2022) when the Omicron BA.1 and BA.2 waves hit Toronto, the C28311T mutation rapidly took over from the wild type, as revealed by nPCR-LC-MS in our previous study (Peng et al. 2022). This C28311T mutation has been consistently detected in all wastewater samples since late Dec 2021 as a universal mutation across all Omicron sub-lineages. In Period III (Aug 1, 2022 - Nov 30, 2022), in addition to the C28311T universal Omicron mutation, an additional mutation, A28330G was also detected in the N1 probe region. The A28330G mutation, with the highest proportion detected on Oct 16, 2022 (77% ± 4, Fig. 2B), was a signature for variant BA.5.2 sub-lineages including BF.7 and BF.10. In Period IV (Dec 1, 2022 - Dec 28, 2022) corresponding to the wave of BQ.1/BQ.1.1, another second additional mutation, C28312T in the N1 probe region, was simultaneously detected in Toronto wastewaters, with the highest proportion (48% ± 1) detected on Dec 26, 2022. Collectively, the nPCR-LC-MS clearly demonstrated the shift of N1 mutation patterns in TAB wastewaters across four time periods corresponding to the occurrence of different variant waves, as shown in Fig. 2.

### 2.3. The shift in N2/N1 ratios is associated with N1 mutation patterns in wastewaters

To investigate the potential relationship between the N2 and N1 discrepancy and N1 mutation patterns, we calculated N2/N1 ratio in wastewaters across the four different periods defined by the mutation patterns confirmed by the nPCR-LC-MS analysis. Recall that the quantified N1 RT-qPCR amplification products were used for the nested-PCR and following LC-MS analysis, thereby eliminating errors introduced in subsampling, and thus ensuring more accurate and precise data to investigate relationships between mutations and RT-qPCR quantifications.

As plotted in Fig. 2C, the N2/N1 ratios varied systematically in the different time periods (I-IV) defined by proportions of mutations. In Period I (yellow region), the N2/N1 ratio was close to the expected value of 1, varied slightly when overall signal strength was very low. There was a very noticeable increase in the N2/N1 ratio at the onset of Period II (green region) corresponding to the single universal Omicron mutation C28311T, and the ratio remained elevated through Period III (blue region) when the second mutation A28330G occurred. In Period IV (purple region), the A28330G mutation located 18 bases away at the probe 3’end from the universal Omicron mutation (C28311T) diminished and an alternate second mutation C28312T next to the universal Omicron mutation closer to the probe 5’ end started to increase, when N2/N1 ratios recovered a little (Fig. 2C).

The overall average N2/N1 ratios were calculated for each period (Fig. 3), clearly showing these differences between periods. The N2/N1 ratios in Period IV (mean = 1.54 ± 0.35, n = 31) with both C28311T and C28312T mutations were significantly higher than those in Period I (mean = 0.84 ± 0.44, n = 275) with no mutation, but not statistically different than Period II (mean = 1.42 ± 0.35, n = 182) with just the Omicron mutation C28311T (*p* = 0.16), suggesting that the secondary mutation C28312T (in addition to C28111T) at the probe 5’ end did not have a large impact on quantification beyond regular Omicron strains. However, in Period III when the second mutation A28330G close to the probe 3’ end occurred, the mean ratio (1.73 ± 0.33, n = 82) was significantly higher than Period II and Period IV, indicating the importance of mutation at the probe 3’ end, together with Omicron universal C28311T, on accuracy of quantification. A similar N1 “dropout” pattern corresponding to various VOC waves was also observed in the surveillance data from fourteen other labs who have been monitoring SARS-CoV-2 level in wastewater in Ontario (Thakali et al. 2023). All these results clearly demonstrated the link between the observed N2/N1 discrepancy and N1 mutation pattern. Nevertheless, wastewater often contained multiple strains of SARS-CoV-2 with a mixture of mutations in the N1 probe region, making it difficult to directly prove the impact of mutation in N1 probe region on N1 RT-qPCR quantification. Therefore, we proceeded to directly measure the impact of these single or double mutations on qPCR quantification by individual DNA oligomers representing these mutations.

**Fig. 3.**
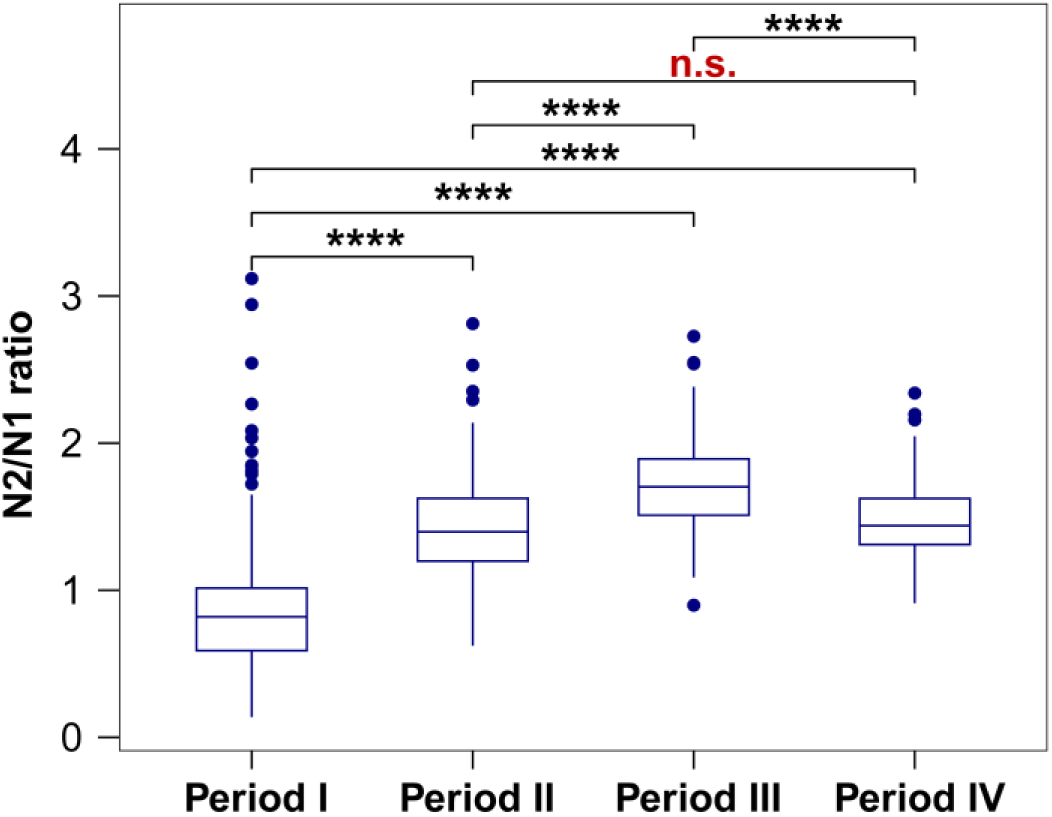
Differences of mean N2/N1 ratios among the four periods. The differences were very significant (****, *p* < 0.0001) when tested by Wilcox among periods I, II, III, and IV, except not significant (n.s., *p* = 0.16) between period II and IV.

### 2.4. Using oligomers containing the signature mutations to confirm underestimation of N1 in qPCR

To further confirm the effect of specific mutations on N1 qPCR quantification, we designed four DNA oligomers which included both the N1 and N2 gene regions (Table S1). Since there was no mutation in the primer binding regions of the CDC N1 and N2 assays, the cDNA synthesis in the reverse transcription step of the mutated strains should not be impacted. Therefore, the underestimation must be associated with the PCR stage, and thus using DNA-based oligomers would mimic that impact. In our design, while the N2 region was identical to the wildtype sequence, the N1 region in these oligomers varied to represent different mutations as shown in Fig. 1 and Table S1. Since these oligomers all shared the same N2 sequence, the CDC N2 qPCR assay calibrated with the wild type ConcatP standard (Fig. S1) was used to quantify the concentrations of these oligomers. The same batch of serially diluted oligomers from N2 assay was used as the template for CDC N1 qPCR assay to examine the impact of the mutations on N1 quantification. All N1 and N2 reactions were conducted on the same PCR plate to minimize error.

The true concentrations of the oligomers calibrated by N2 qPCR standard curve were compared to the estimations based on N1 qPCR using ConcatP as standard (Fig. 4). Delayed Ct values were observed in N1 variant oligomers, and the resulting concentrations determined using the wild type (ConcatP) standard curve are therefore lower. As shown in Fig. 4, the estimated N1 concentrations of the oligomers with mutations (i.e., Omicron, BQ.1*, BA.5.2 and BF.10) were all lower compared to the true concentrations. In general, for the serially diluted standards, the N1 quantifications for oligomer representing general Omicron with single mutation by ConcatP standard curve were 1.53-fold lower than true value. The N1 quantification for oligomer corresponding to BA.5.2 with two mutations including one at the probe 3’ end had the most effect which were 2.28-fold lower than true value, following BF.10 corresponded three mutations with 2.18-fold lower than true value, and BQ.1* corresponded two mutations with 1.65-fold lower than true value.

**Fig. 4.**
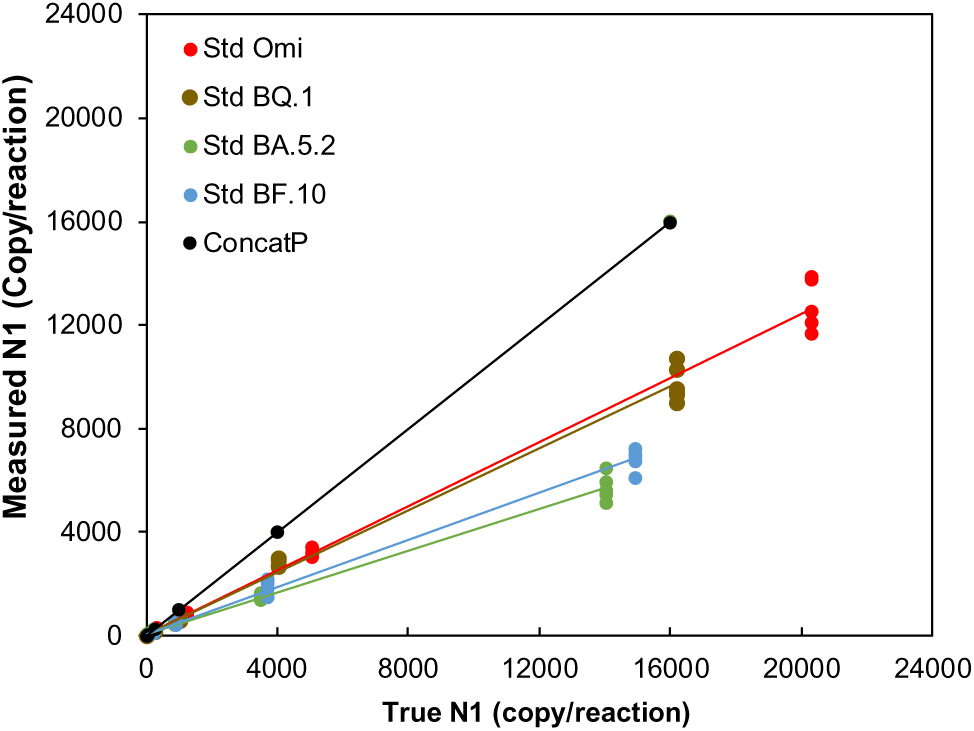
Measured vs true N1 copies per reaction as a function of sequence. True concentrations of each oligomer (measured via the N2 reaction) are plotted on the X-axis versus their corresponding measured value on the Y-axis calibrated using the wildtype sequence (ConcatP) as standard. Black line shows wild type quantification and a perfect match between measured and true values. For all targets with mutations, quantification is underestimated, which is especially pronounced for BA.5.2 (green) and BF.10 (blue) where the measured value is about half the true value. The measured values for BQ.1 (brown), and all Omicron (red) are about 25% depressed.

The oligomer experiments also revealed that the largest deviation occurred with the oligo containing both C28311T and A28330G mutations, consistent with the largest discrepancy between N2 and N1 observed in TAB wastewater during the BA.5.2 wave. The close quantifications between BQ.1 oligomer and Omicron universal oligomer also match the wastewater results where no significance was observed between period II and IV. Therefore, the oligomer experiments and the shifts in the ratio of N2/N1 in the monitored TAB wastewaters, along with the proportions of mutations in the actual TAB samples inferred by LC-MS, confirm that the mutation in the N1 probe region was directly associated with the underestimation of the N1 quantification in RT-qPCR when using the wildtype sequence for calibration.

### 2.5. Implications for utility of the CDC N1 RT-qPCR assay

The effect of primer/probe-template mismatch on quantification in qPCR has been investigated in other targets for SARS-CoV-2 (Bozidis et al. 2022, Hasan et al. 2021). Especially, mutation in the probe binding region was found to largely impact qPCR amplification by delaying amplification or causing failure, which were also used to design mutation/variant detection methods such as the N200 assays to quantify Delta/Omicron/Universal SARS2 (Fuzzen et al. 2023), the D3Ll assay to detect Alpha variant (Graber et al. 2021), and the various assays targeting the S gene to measure Alpha/Beta/Gamma variants (Liu et al. 2023, Peterson et al. 2022, Vega-Magaña et al. 2021). N gene dropout or failure was also reported using some commercial SASR2 RT-qPCR kits when mutations were found in the N200 region (28881-28883, 28896-28898, in B.1.1.318 variant) by sequencing (Bozidis et al. 2022). Our study also demonstrated that both the number of mutation (e.g., 0 vs. 1 mutation vs. 2 mutations) and the mutation position on the probe binding region (e.g., the second mutation on the probe 3’ end vs. 5’ end) influence the amplification of the N1 target, and thus affected the degree of underestimation when using the wildtype N1 sequence as calibration standards.

Although there were up to three mismatches between the CDC N1 probe and the Omicron strains that we examined in this study, all templates were still amplified. The problem with the N1 underestimation with these mutated strains was that the standardized materials used in our routine analysis contained the wildtype sequence that didn’t have the delayed amplification as the mutated strains. In other words, the amplification behavior in our standardized materials was not comparable to the actual TAB wastewater samples or oligomers with mutations. When we ran serial dilutions of the N1 variant oligomers and ConcatP in the CDC N1 qPCR, we still achieved relatively good amplification efficiency (97.9-102.2%) for the standard curves although the y-intercepts were delayed in the oligomers with mutations (Fig. S1).

Given that the templates could still be amplified in the CDC N1 qPCR reaction despite the mismatches, there are multiple ways to tune the assay to achieve more accurate N1 quantification. One adjustment would be to switch to a different standardized material to better represent the N1 probe region mutation profile in the actual wastewater samples. When changing to use the oligomer representing the dominant mutation type in each period as qPCR standards instead of using the wildtype ConcatP, an N2/N1 ratio was closer to 1 with the newly calibrated N1 concentrations (Fig. S2). Previously this was difficult to execute with multiple strains co-existing in wastewaters with different mutation types. It might be feasible now, as XBB has been the sole main type of SARS-COV-2 variant (Public Health Ontario, 2023) since early 2023 and all sub-lineages of XBB only have the C28311T mutation in the N1 probe region. Another modification to improve the CDC N1 qPCR assay was to run the N1 assay at a lower annealing temperature to reduce the thermal barrier of probe-template binding and minimize Ct delay. A third option is to redesign the N1 probe to bind to a more conserved region within the N1 gene, or switch to a different assay target that is less prompt to mutation such as the N200 (Fuzzen et al. 2023).

Another approach to increase the N1 CDC assay accuracy is to switch from qPCR platform to digital PCR platform. Digital PCR does not rely on amplification curves, i.e, quantification only depends on the positive and negative partition counts at the end of the run instead of relying on amplification behavior and dynamic. Since all mutated strains are still amplifiable in the N1 CDC PCR reaction, mutation only changes the fluorescent amplitude but not the positive vs. negative partition counts (Fig. S3, Table S5). As proof of concept, we selected two TAB samples at different dates to represent each period and conducted CDC N1 reaction on RT-dPCR. The N2/N1 ratios from RT-dPCR were all close to 1 (ranged from 0.8 to 1.14) in samples from all different periods (Table S6). Therefore, switching to dPCR may be one option for accurate quantification of SARS-CoV-2 in wastewater in the future, although a cost/benefit analysis would be required, as dPCR is currently quite a bit more expensive per sample (approximately CAD $4 per RT-qPCR single-plex reaction, while CAD $12 per RT-dPCR single-plex reaction).

## 3. Materials and Method

### 3.1. Samples and Reagents

Ashbridges Bay (TAB) wastewater treatment plant is Canada’s biggest wastewater treatment plant serving over 1.5 million people (Pileggi et al. 2022). Post-grit composite influent samples (24-hour) from TAB were collected three to five times per week since Feb 11, 2021 (Peng et al. 2022, Pileggi et al. 2022). 500 mL – 1 L subsamples were collected onsite and were stored at 4 °C and shipped as soon as possible, typically on the same day, to the laboratory at the University of Toronto in a cooler on ice.

All plasticware (tips, tubes, plates, etc.) used in extraction and PCR were DNase- and RNase-free. Probes for N1 (cat. 10006832) and N2 (cat. 10006835) were purchased from Integrated DNA Technologies (IDT). Two PCR standards were purchased: the N-plasmid control from IDT (10006625) and an in-house designed concatenated plasmid (ConcatP, GenBank accession number OR994921) purchased from Twist, both contain the complete wild-type sequence spanning the N1 and N2 regions. Four N1 variant oligomers were designed based on the consensus N1 sequences of each group of BA.1, BQ.1, BF.7 and BF.10 randomly selected from the corresponding lineage in the GISAID database. Each N1 variant oligomer contained the CDC N1 region of the corresponding variant group with their specific mutation(s) as well as the original wild type CDC N2 region. All the four N1 variant oligomers (177 bp) were then purchased from Eurofins; exact sequences are provided in Table S1. Other primers and shorter oligomer standards (<40 bp) required for LC-MS were purchased from Invitrogen, Thermo Fisher Scientific; their sequences are provided in Tables S3 and S4). Methanol was purchased from Thermo Fisher Scientific. Triethylamine (TEA, 471283) and hexafluoroisopropanol (HFIP, 105228) were obtained from Sigma-Aldrich. Unless otherwise specified, all other reagents were analytical grade.

### 3.2. Viral RNA extraction from wastewaters

TAB wastewater samples (500 mL - 1 L) were shipped to the lab three times per week and RNA extraction started immediately upon sample arrival. After being well mixed, 80 mL were sub-sampled from the original shipment bottle and split into two 50 mL falcon centrifuge tubes (40mL each). After centrifugation at 10,000 × g for 45 minutes at 4°C, pellets from the two Falcon tubes were resuspended with 1 - 2 mL remaining wastewater and transferred to a single 2 mL microcentrifuge tube. The pooled wet pellet was centrifuged at 13,000 × g for one minute and the supernatant was removed. The pellet wet mass was recorded. RNA was extracted from the pooled pellet using Qiagen’s RNeasy PowerMicrobiome Kit (Qiagen, Germantown, MD), with 10 μL beta-mercaptoethanol (Bioshop, MER002) and 100 μL phenol: chloroform: isoamyl alcohol (25: 24: 1, v/v) solutions (Invitrogen, CAT# 15593031, USA) added to the bead beating tube along with the lysis buffer from the kit. DNase step was skipped. All RNA extracts were eluted in a volume of 100 μL. A whole-process-control was included in each batch of extractions that contained all the same components except the sample.

### 3.3. Reverse transcriptase qPCR and digital PCR assays for N1 and N2

The published CDC assays (Lu et al. 2020), targeting at N1 and N2 gene regions, were used for the routine quantification of the SARS-CoV-2 viral signal in wastewaters. In brief, each RT-qPCR reaction contained 2.5 μL 4 × TaqMan Fast Virus 1-Step Master Mix (CAT# 4444436, ThermoFisher Scientific), 125 nM probe and 500 nM each of forward and reverse primers, 4 μL of RNA sample or standard and water to a final reaction volume of 10 μL. Bio-Rad CFX Opus 384 Real-Time PCR System was applied for all RT-qPCR assays. For routine wastewater qPCR analyses, the linearized N-plasmid (from Feb 11, 2021 to Aug 08, 2021) or the linearized ConcatP (from Aug 09, 2021 to Dec 29, 2022) were used as standards. Calibration curves for both N1 and N2 were included on each plate. All standards for RT-qPCR, including IDT N-plasmid, ConcatP and the four variant N1 oligomers, were diluted in a polyA-TE (Tris and EDTA) carrier matrix as described SI.

RT-digital PCR (RT-dPCR) was performed in 40-µL reaction mixtures using the QIAcuity OneStep Advanced probe Kit (Cat No. 250132) (Ahmed et al. 2022). Each RT-dPCR reaction contained 10 µL of 4x Probe Master Mix, 0.4 µL 100x RT Mix, 0.4 µM each of forward and reverse primers, 0.2 µM probe, 5 µL of template RNA and PCR-grade water to a final volume of 40 µL. The reactions were prepared in 200 µL PCR tubes, and then loaded on to 26K 24-wells (QIAGEN) after well mix. The Nanoplate was then loaded onto the QIAcuity dPCR 5-plex platform (Qiagen) and subjected to a workflow that included: (i) a priming step to create the nano-size partitions; (ii) an amplification/thermocycling step (RT at 50°C for 30 min, initial denaturation at 95°C for 2 minutes, and 45 cycles of 5 second at 95 °C and 30 second at 60°C); and (iii) a final imaging step in the FAM channel. A no-template control and a positive control were also included in each RT-dPCR run. Data were analyzed using the QIAcuity Suite Software V2.1.8.23 (Qiagen, Germany). Threshold lines to separate the positive and negative partitions were set manually to be consistent among the reactions on the same plate, and the quantities exported as gene copies/µL of reaction (Table S5).

All RNA extracts were run in technical triplicates for RT-qPCR and technical duplicates for RT-dPCR. The limit of detection (LOD) for qPCR was defined at a theoretical lowest number (1 copy) in each reaction (0.31 copies per mL of wastewater). The limit of quantification (LOQ) of RT-qPCR was set at the lowest concentration of standard curves which was 3.9 copies per reaction (1.25 copies per mL of wastewater) for both N1 and N2. The quantification for low template concentration is not reliable, and in this study, all results with copies per reaction lower than half of LOD were replaced with 0.5 copies/reaction (0.16 copies per mL of wastewater). The LOD for RT-dPCR was approximately 2 copies per reaction (i.e., ∼0.05 copies per µL rection).

### 3.4. nPCR-LC-MS Method

The nested polymerase chain reaction and liquid chromatography-mass spectrometry (nPCR-LC-MS) method developed and reported previously (Peng et al. 2022), with modification, was employed to detect and quantify mutations in wastewater samples. In brief, the resulting DNA products amplified in the CDC-N1 RT-qPCR assay were diluted 100-fold in water and used as templates for nested PCR. This procedure ensured that the proportions of mutation(s) measured by LC-MS were from the same RNA template as quantified by qPCR. While the nested PCR (nPCR) primers for the amplification of 28310 – 28312 region were the same as reported (Peng et al. 2022), another pair of nPCR primers were designed for the amplification of 28328 – 28330 region in this study (Table S3 and Table S4). Chromatograms for 28310 – 28312 regions from samples and standards were shown in Fig. S4. The final nested PCR product (50 μL) was digested with sequencing grade trypsin (V5113, Promega) overnight, and then precipitated by adding ethanol to 70% and stored at −80°C for 1 hour. After centrifuge at 4 °C for 30 min, the supernatant was removed and the pelleted DNA was re-dissolved in water. Subsequently, it was transferred to a 96-well plate for sample loading by the Vanquish UPLC system (Thermo Scientific). The separation was conducted in a ACQUITY UPLC Oligonucleotide BEH C18 Column (130Å, 1.7 µm, 2.1 mm X 50 mm, Waters, SKU: 186003949) with mobile phase A (water with 15 mM TEA and 25 mM HFIP) and mobile phase B (methanol) at a flow rate 0.2 mL/min with a gradient elution starting at 5% B for 2 min, then increasing from 5 to 30% B over 7 min, and then increasing to 99% B over 6.5 min. Finally, the eluent was returned to 5% B in 0.5 min, and held at 5% B for 2 min prior to next run. The molecular identification was performed on a Q-Exactive orbitrap. A spray voltage of 3.0 kV and an ion transfer tube temperature of 300 °C were used for ionization and desolvation. Precursor spectra were acquired from m/z 1000 to 5000 in negative ionization mode at a resolution of 140,000.

More details about RNA extraction, nPCR and RT-qPCR are provided in SI.

### 3.5. Data analysis

The N2/N1 ratio for each sample was calculated using the average of the technical triplicate quantifications in RT-qPCR. A fourteen-day moving average of the N2/N1 ratio was calculated and plotted using Microsoft excel software. Since results with lower than half of RT-qPCR LOD were replaced with 0.5 copies/reaction for both N1 and N2, samples where 2 or 3 of the technical triplicates equal to 0.5 copies/reaction were excluded in the N2/N1 ratio trend analysis to ensure that only data with high quantification certainty were included in our study. The proportion of each mutation was calculated as the abundance of the target mutation divided by the sum of the abundance of all genotypes detected in the nPCR amplification product. The cut-off dates for each period were defined as the earliest date when the proportion of the dominant mutation reached half of its highest observed proportion at our monitored site (i.e., TAB). The significance of the differences of N2/N1 ratios among the 4 periods were analyzed using the Willcox test.

## Data Availability

All data produced in the present study are available upon reasonable request to the authors

## Supporting Information Available

The supporting information provides tables and figures addressing: (1) Detailed information for RNA extraction and RT-qPCR, Mass Spectrometry detection of mutations in wastewater, and preparation of DNA oligomers with variant N1 sequences; (2) tables for sequences of N1-N2 DNA oligomers, GISAID numbers for N1 sequence alignment, sequences of primers and probes used in this study, sequences and *m/z* for short oligomers, and MIQE check lists for qPCR and dPCR; (3) figures for N1 and N2 standard curves, N2/N1 ratio when N1 calibrated by standard curves with corresponding mutations, N2/N1 ratio comparison analyzed by qPCR and dPCR, and chromatograms of sense (SS) and anti-sense strands (AS) of nPCR amplicons containing A28330G region or oligonucleotide standards.

## Acknowledgements

This research was supported by the Ontario Wastewater Surveillance Initiative, and the Natural Sciences and Engineering Research Council (NSERC) Alliance Grant (ALLRP 576140). The authors acknowledge the support of instrumentation grants from the Canada Foundation for Innovation and the Ontario Research Fund.

