## Supplemental materials for "Underestimation of SARS-CoV-2 in wastewater due to single or double mutations in the N1 qPCR probe binding region"

Number of pages: 15

Number of tables: 6

Number of figures: 4

*Details about RNA extraction and RT-qPCR*

All wet weight of pellets from 80 mL of TAB wastewater did not exceed the sample limit of Qiagen's RNeasy PowerMicrobiome Kit (Qiagen, Germantown, MD), which is 250 mg. Total nucleotide concentrations (median = 1574 ng/μL) were quantified by NanoDrop, and RNA extracts with A260/A280 at about 2.1, and A260/A230 at about 2.4 were applied in RT-qPCR reaction. Extracted RNA samples were all stored at -80 °C. Two positive RNA controls (high and low), which were extracted from wastewater samples, were split and stored at -80 °C for daily use.

The N-plasmid and ConcatP were both transformed into DH5Alpha *E.coli* strains. Single strain was selected, and plasmid was extracted from 500 mL LB culture by HiSpeed Plasmid Midi Kit (Qiagen). After sanger sequencing (Sickkids), the plasmids were digested with Pcil (NEB) overnight and then purified by QIAquick PCR Purification kit (Qiagen). The purity check was conducted by nanodrop and bioanalyzer, then quantified by Qubit (Agilent) and latter by dPCR.

Singleplex RT-qPCR quantification of the SARS-CoV-2 viral signal was performed by targeting the N1 and N2 gene regions. Another target for Pepper mild mottle virus (PMMoV) (Rosario et al. 2009) was also included on each RT-qPCR plate, which were loaded at original concentration of RNA and 10-fold dilution for each sample as inhibition check. Ct values were compared, and inhibition is called when more than one Ct shift (deltaCt less than 2.3 as expected difference is 3.3) was observed between original and 10-fold diluted RNA. Fortunately, no TAB RNA showed inhibition in this study. The PCR conditions for RT-qPCR were: RT at 50°C for 5 min, 95°C for 20 sec, 45 cycles of 95°C for 3 sec followed by 60°C for 30 sec. Standard curves for each target, three negative controls in technical duplicates (no template control, whole process control, and polyA carrier control), and two positive controls in technical triplicates (with concentrations similar to the tested samples) were run along with samples on each 384 PCR clear plate. Quantification was performed via 7-point ConcatP standard curves include a 4-fold serial dilution with concentrations from 3.9 to 1.6 × $10^4$ copies/reaction for both N1 and N2. Threshold for all RT-qPCR files were set at 100 (within the linear range of the amplification curve) to easier compare Ct reproducibility among plates. All standard curves for routine analysis were linear ($R^2$ ⩾ 0.99) and the primers efficiency were between 95% – 105% for both N1 and N2.

*Details of Mass Spectrometry detection of mutations in wastewater*

Before and after opening the N1 qPCR amplification products, the working bench, pipettes, tips boxes, ect. need to be wiped with 1% bleach followed by 70% ethanol, and UV treated for 30 min to avoid contamination. All the three technical triplex amplicons from N1 qPCR for each sample were 100-fold diluted in waster and loaded into three nPCR reactions. Three negative controls, one from each negative controls of PolyA carrier, WPB, and NTC on N1 qPCR plate were also 100-fold diluted and included for each batch of nPCR. The reaction for nested PCR contains 500 nM of the forward and reverse primers were mixed with 25 µL Taq 2× Master Mix. The mixture (final volumn 50 µL) was heated at 95 °C for 5 min, 30 cycles of 95 °C for 30 s, 46 °C for 30 s, and 68 °C for 10 s, and finally 68 °C for 2 min. The final nested PCR products were digested with sequencing grade trypsin for overnight, then added 5 µL of sodium acetate (3 M, pH 5.3) and 125 µL of cold ethanol. After vertexing, the samples were stored at −80 °C for one hour, then centrifuged at 4 °C for 30min. The resulting pellet was resuspended in 50 µL water. The purified DNA was transferred to a 96-well plate for sample loading by the Vanquish UPLC system (Thermo Scientific).

*Preparation of DNA oligomers with variant N1 sequences*

All DNA oligomers were diluted in polyA as carrier matrix based on the mass weight provided by the manufacturer to a concentration close to $4 \times 10^3$ copies/μL, then conducted 4-fold serial dilution until about 1 copy/μL. Four μL of all the 7 serially diluted standards for the four variant N1 oligomers were loaded into N1 reactions, and the highest two concentrations ($4 \times 10^3$ and $1 \times 10^3$ copies/μL) were loaded into N2 reactions for quantification. On the same plate, wildtype ConcatP 7-point standard curves (from $4 \times 10^3$ copies/μL to 1 copies/μL) were also included for both N1 and N2 reactions. Different from routine analysis, for all the oligomers and ConcatP, each concentration was run with 6 replicates, except that the lowest concentration was run with 9 replicates. All variant N1 oligomers were quantified by the ConcatP N2 calibrate curve on this plate. After quantified, standard curves for N1 with different mutations were generated based on the quantification from N2 reaction and Ct value in N1 reaction.

Table S1. Oligomer Sequences (5’ to 3’; 177 bp) used in PCR reactions corresponding variants in the N1 region. Bases in red show mutations compared to wild type sequence, bases in shade show N1 probe binding region.

| | |
|---|---|
| Omicron Universal | TGTCTGATAATGGACCCCAAAATCAGCGAAATGCACTCCGCATTA<br>CGTTTGGTGGACCCTCAGATTCAACTGGCAGTAACCAGAATGAA<br>CTGATTACAAACATTGGCCGCAAATTGCACAATTTGCCCCCAGCG<br>CTTCAGCGTTCTTCGGAATGTCGCGCATTGGCATGGAAGTCAC |
| BA.5.2 | TGTCTGATAATGGACCCCAAAATCAGCGAAATGCACTCCGCATTA<br>CGTTTGGTGGGCCCTCAGATTCAACTGGCAGTAACCAGAATTAAC<br>TGATTACAAACATTGGCCGCAAATTGCACAATTTGCCCCCAGCGC<br>TTCAGCGTTCTTCGGAATGTCGCGCATTGGCATGGAAGTCAC |
| BQ.1 | TGTCTGATAATGGACCCCAAAATCAGCGAAATGCACTTCGCATTA<br>CGTTTGGTGGACCCTCAGATTCAACTGGCAGTAACCAGAATGAA<br>CTGATTACAAACATTGGCCGCAAATTGCACAATTTGCCCCCAGCG<br>CTTCAGCGTTCTTCGGAATGTCGCGCATTGGCATGGAAGTCAC |
| BF.10 | TGTCTGATAATGGACCCCAAAATCAGCGAAATGCATTCCGCATTA<br>CGTTTGGTGGGCCCTCAGATTCAACTGGCAGTAACCAGAATGAA<br>CTGATTACAAACATTGGCCGCAAATTGCACAATTTGCCCCCAGCG<br>CTTCAGCGTTCTTCGGAATGTCGCGCATTGGCATGGAAGTCAC |

85 Table S2. Sequences used to identify common mutations in variant subgroups

| Main group | Subgroup included | GISAID ID or NCBI accession # |
|---|---|---|
| Wild type | | WT_MN908947.3 |
| Alpha | | EPI_ISL_710528 |
| Omicron with one mutation | BA.1, BA.2, BA.2.75, XBB | EPI_ISL_14758997, EPI_ISL_15115194, EPI_ISL_15115244, EPI_ISL_6841980, EPI_ISL_7190366, EPI_ISL_15090226, EPI_ISL_15321209, EPI_ISL_15392552, EPI_ISL_16242475, EPI_ISL_16322905, EPI_ISL_16323406 |
| BF.7 (and other BA.5.2.1) | | EPI_ISL_14314630, EPI_ISL_14314630, EPI_ISL_15054483, EPI_ISL_15071770, EPI_ISL_16086537 |
| BF.10 | | EPI_ISL_14314630, EPI_ISL_15054741 |
| BQ.1 and BQ.1.1 | | EPI_ISL_15415930, EPI_ISL_15046698, EPI_ISL_15101240, EPI_ISL_15127947 |

Table S3. Sequences of primers and probes for N1 and N2 RT-qPCR, and nested PCR primers for nPCR-LC-MS analyses

| Primer/Probe | Sequence (5'-3') | Amplicon size | Reference |
|---|---|---|---|
| 2019-nCoV_N1 forward primer | GACCCCAAAATCAGCGAAAT | | |
| 2019-nCoV_N1 reverse primer | TCTGGTTACTGCCAGTTGAATCTG | | |
| 2019-nCoV_N1 probe (IDT) | 6-FAM-ACC CCG CAT/ZEN/ TAC GTT TGG TGG ACC-IOWA BLACK FQ | 73bp | U.S. CDC |
| 2019-nCoV_N2 forward primer | TTA CAA ACA TTG GCC GCA AA | | |
| 2019-nCoV_N2 reverse primer | GCG CGA CAT TCC GAA GAA | | |
| 2019-nCoV_N2 probe (IDT) | 6-FAM-ACA ATT TGC/ZEN/CCC CAG CGC TTC AG-IOWA BLACK FQ | 68bp | U.S. CDC |
| P13 nest forward primer | TCAGCGAAATGCA | | Environ. Sci. Technol. |
| P13 nest reverse primer | CCAAACGTAATGCG | 31bp | Lett. 2022, 9, 7, 638–644 |
| Nested 28330 Primer F | CGCATTACGTTTGGT | | |
| Nested 28330 Primer R | CAGTTGAATCTGAGGG | 35bp | This study |

Table S4. Predicted sequences for nested PCR products and corresponding mass to charge ratio (m/z) for analysis by mass spectrometry.

| Target | Amplicon Sequence (5'-3') | m/z |
|---|---|---|
| P13 amplicon (Wildtype) | TCAGCGAAATGCACCCCGCATTACGTTTGGa* | 2368.8# |
| P13 amplicon (Omicron Universal) | TCAGCGAAATGCACTCCGCATTACGTTTGGa | 2372.55# |
| P13 amplicon (BQ.1) | TCAGCGAAATGCACTTCGCATTACGTTTGGa | 2376.31 |
| A28330 amplicon (WT, BQ.1) | CGCATTACGTTTGGTGGACCCTCAGATTCAACTGa | 2679.45 |
| A28330G amplicon (BA.5.2) | CGCATTACGTTTGGTGGGCCCTCAGATTCAACTGa | 2683.20 |
| P13 amplicon (Wildtype) anti-sense | CCAAACGTAATGCGGGGTGCATTTCGCTGAa | 2388.81# |
| P13 amplicon (Omicron Universal) anti-sense | CCAAACGTAATGCGGAGTGCATTTCGCTGAa | 2384.81# |
| P13 amplicon (BQ.1) anti-sense | CCAAACGTAATGCGAAGTGCATTTCGCTGAa | 2380.81 |
| A28330 amplicon (WT, BQ.1) anti-sense | CAGTTGAATCTGAGGGTCCACCAAACGTAATGCGa | 2696.20 |
| A28330G amplicon (BA.5.2) anti-sense | CAGTTGAATCTGAGGGCCCACCAAACGTAATGCGa | 2692.20 |

* Taq polymerase has nontemplate-dependent terminal transferase activity that adds a single deoxyadenosine (denoted a) to the 3´-ends of PCR products.

### Same as citation (Peng et al. 2022)

**Table S5.** dPCR quantifications exported from QIAcuity Suite Software V2.1.8.23

| Sample date | target | Channel | Partition (valid) | Partition (positive) | Partition (negative) | Threshold | CI (95%) | Concentration (copies/µL) |
|---|---|---|---|---|---|---|---|---|
| Apr-06-21 | N1 | GREEN | 25367 | 35 | 25332 | 40 | 0.331 | 1.825 |
| Apr-06-21 | N1 | GREEN | 25381 | 26 | 25355 | 40 | 0.384 | 1.369 |
| Apr-06-21 | N2 | GREEN | 25375 | 32 | 25343 | 40 | 0.346 | 1.706 |
| Apr-06-21 | N2 | GREEN | 25293 | 27 | 25266 | 40 | 0.377 | 1.45 |
| Apr-28-21 | N1 | GREEN | 22701 | 39 | 22662 | 40 | 0.314 | 2.351 |
| Apr-28-21 | N1 | GREEN | 25405 | 42 | 25363 | 40 | 0.302 | 2.254 |
| Apr-28-21 | N2 | GREEN | 25434 | 34 | 25400 | 40 | 0.336 | 1.811 |
| Apr-28-21 | N2 | GREEN | 22764 | 32 | 22732 | 40 | 0.346 | 1.867 |
| Jan-23-22 | N1 | GREEN | 25349 | 83 | 25266 | 40 | 0.215 | 4.371 |
| Jan-23-22 | N1 | GREEN | 25328 | 64 | 25264 | 40 | 0.245 | 3.472 |
| Jan-23-22 | N2 | GREEN | 25407 | 58 | 25349 | 40 | 0.257 | 3.202 |
| Jan-23-22 | N2 | GREEN | 25397 | 77 | 25320 | 40 | 0.223 | 4.306 |
| Jan-27-22 | N1 | GREEN | 25089 | 19 | 25070 | 40 | 0.45 | 1.065 |
| Jan-27-22 | N1 | GREEN | 25292 | 33 | 25259 | 40 | 0.341 | 1.832 |
| Jan-27-22 | N2 | GREEN | 25306 | 28 | 25278 | 40 | 0.37 | 1.537 |
| Jan-27-22 | N2 | GREEN | 25374 | 32 | 25342 | 40 | 0.346 | 1.702 |
| Oct-16-22 | N1 | GREEN | 25439 | 119 | 25320 | 40 | 0.18 | 6.263 |
| Oct-16-22 | N1 | GREEN | 25443 | 135 | 25308 | 40 | 0.169 | 7.2 |
| Oct-16-22 | N2 | GREEN | 25402 | 117 | 25285 | 40 | 0.181 | 6.303 |
| Oct-16-22 | N2 | GREEN | 25364 | 113 | 25251 | 40 | 0.184 | 6.189 |
| Oct-23-22* | N1 | GREEN | 25380 | 43 | 25337 | 40 | 0.299 | 2.328 |
| Oct-23-22* | N1 | GREEN | 25331 | 49 | 25282 | 40 | 0.28 | 2.701 |
| Oct-23-22* | N2 | GREEN | 25436 | 59 | 25377 | 40 | 0.255 | 3.158 |
| Oct-23-22* | N2 | GREEN | 25234 | 48 | 25186 | 40 | 0.283 | 2.567 |
| Dec-26-22 | N1 | GREEN | 25364 | 140 | 25224 | 40 | 0.166 | 7.316 |
| Dec-26-22 | N1 | GREEN | 25425 | 125 | 25300 | 40 | 0.175 | 6.582 |
| Dec-26-22 | N2 | GREEN | 25371 | 132 | 25239 | 40 | 0.171 | 7.051 |
| Dec-26-22 | N2 | GREEN | 25443 | 134 | 25309 | 40 | 0.169 | 7.167 |
| Dec-28-22 | N1 | GREEN | 25392 | 173 | 25219 | 40 | 0.149 | 9.348 |
| Dec-28-22 | N1 | GREEN | 25408 | 156 | 25252 | 40 | 0.157 | 8.391 |
| Dec-28-22 | N2 | GREEN | 25301 | 146 | 25155 | 40 | 0.162 | 7.833 |
| Dec-28-22 | N2 | GREEN | 25427 | 143 | 25284 | 40 | 0.164 | 7.486 |

*RNA sampled on Oct 23, 2022 were run after 3 fold dilution.

Table S6. Comparison of N1 and N2 concentrations (copies/mL wastewater) quantified by RT-qPCR and RT-dPCR. Quantification from RT-qPCR were calibrated by wildtype ConcatP standard. Two TAB samples were selected from each period. Concentrations were shown in average ± STDEV of technical triplicates in RT-qPCR and technical duplicates for RT-dPCR.

| Sample date | Period | Quantified by RT-qPCR | | | Quantified by RT-dPCR | | |
|---|---|---|---|---|---|---|---|
| | | N2 | N1 | N2/N1 | N2 | N1 | N2/N1 |
| 6-Apr-21 | I (no mutation) | 21 ± 3 | 25.9 ± 4.8 | 0.81 | 15.8 ± 1.8 | 16 ± 3.2 | 0.99 |
| 28-Apr-21 | | 32.3 ± 5.4 | 27.1 ± 0.7 | 1.19 | 18.4 ± 0.4 | 23 ± 0.7 | 0.80 |
| 23-Jan-22 | II (single mutation) | 34.7 ± 4.1 | 21.1 ± 3.5 | 1.64 | 37.5 ± 7.8 | 39.2 ± 6.4 | 0.96 |
| 27-Jan-22 | | 15.4 ± 1.3 | 8.7 ± 1.1 | 1.77 | 16.2 ± 1.2 | 14.5 ± 5.4 | 1.12 |
| 16-Oct-22 | III (two mutations, one at 3' end) | 91.4 ± 10.9 | 33.4 ± 1.2 | 2.74 | 62.5 ± 0.8 | 67.3 ± 6.6 | 0.93 |
| 23-Oct-22 | | 100 ± 8.6 | 59.7 ± 10.4 | 1.68 | 28.6 ± 4.2 | 25.1 ± 2.6 | 1.14 |
| 26-Dec-22 | IV (two mutations, both at 5' end) | 110.1 ± 17.2 | 68 ± 3.1 | 1.62 | 71.1 ± 0.8 | 69.5 ± 5.2 | 1.02 |
| 28-Dec-22 | | 92.7 ± 18.5 | 59.2 ± 5.5 | 1.57 | 76.6 ± 2.5 | 88.7 ± 6.8 | 0.86 |

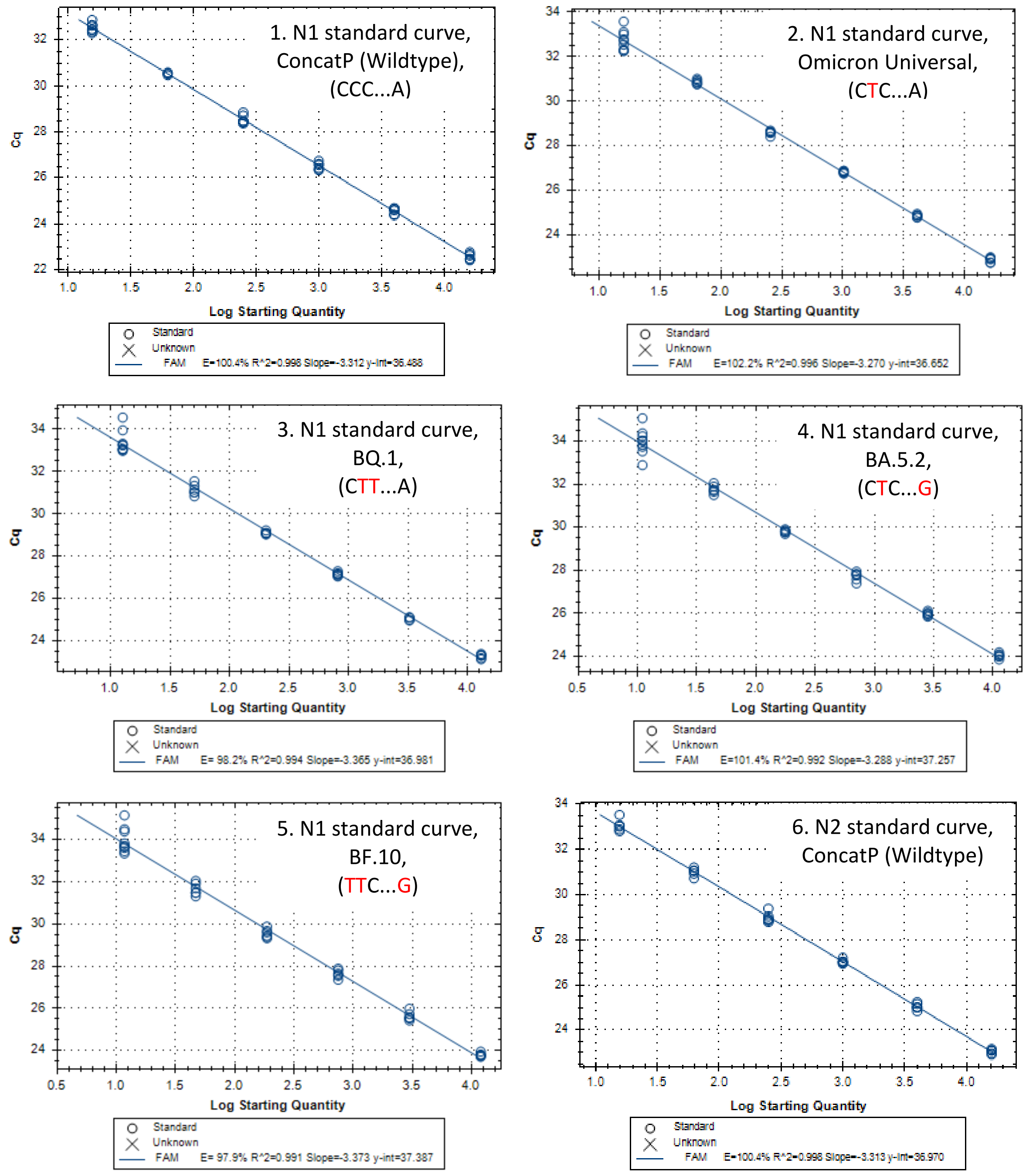


Fig. S1. N1 Calibration curves for wildtype standard (S1.1, ConcatP) and standards incorporating different mutations (S1.2 – S1.5) and N2 wildtype standard curve for quantification of the oligomers (S1.6). Standard curves for N1 reaction ranged from about 1.6E4 copies/reaction varied based on the quantification by N2, with 4-fold serial dilution in polyA carrier until about 16 copies/reaction. All concentrations were run with 6 technical replicates except that the lowest concentration where 9 technical replicates were included.

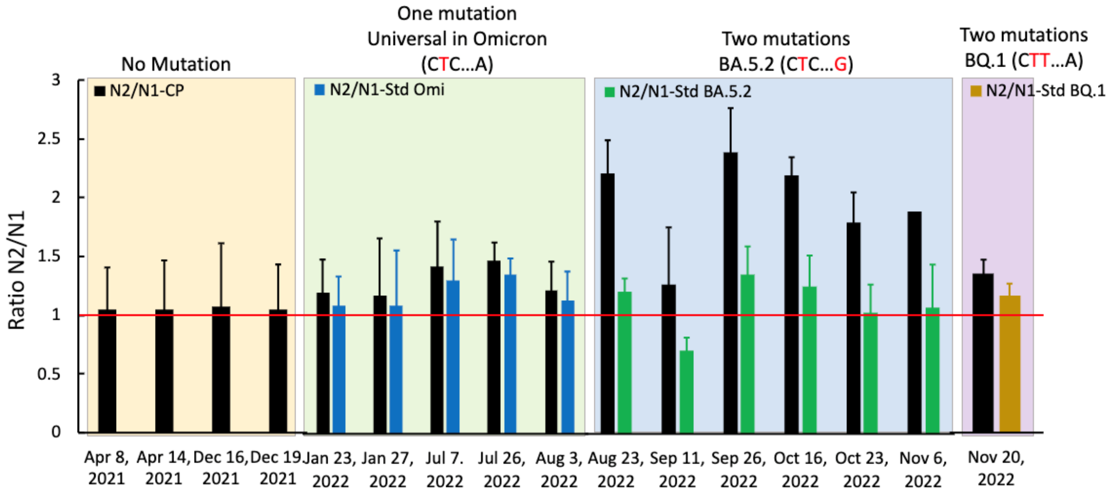


Fig. S2. N2/N1 ratios in different periods. N2/N1 ratios where N1 were quantified using wildtype ConcatP calibration curve were shown in black, and where N1 were quantified using corresponding mutation oligomers were shown in blue (period II), green (Period III), and grey (Period IV). Error bars represent the standard deviation from three technical replicates.

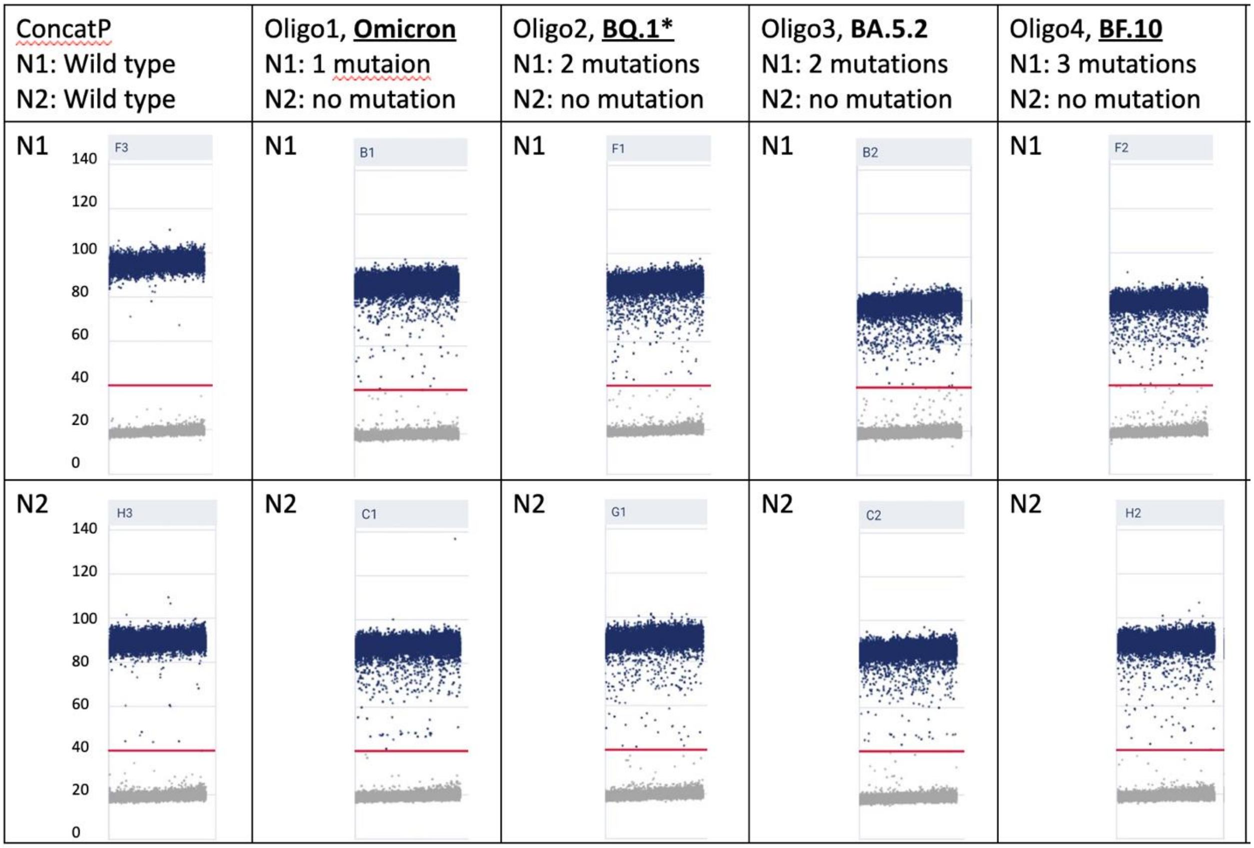


Fig. S3. dPCR 1-D scatter plot to show the fluorescence intensity (blue: positive partitioning; grey: negative partitioning; red line: threshold line manually set at 40)

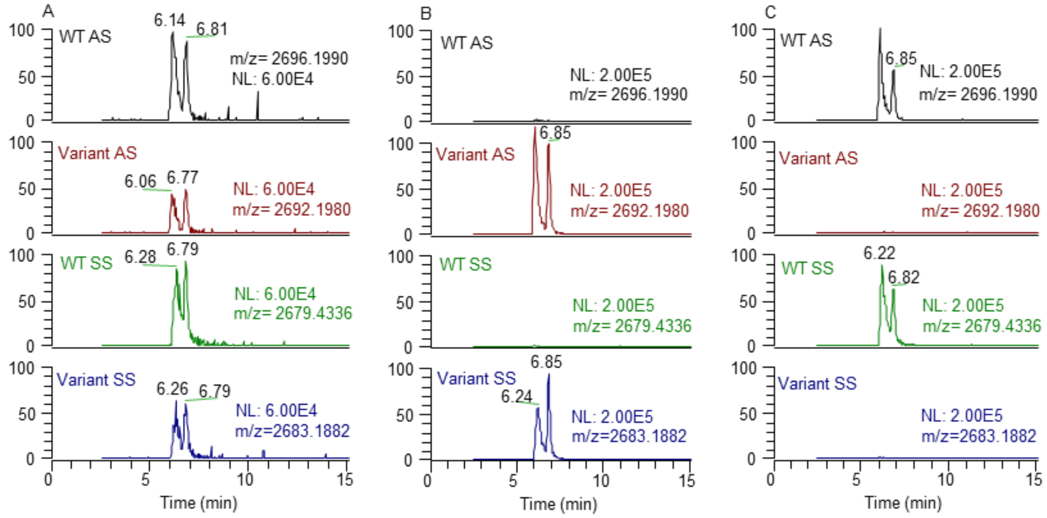


Fig. S4. Chromatograms of sense (SS) and anti-sense strands (AS) of nPCR amplicons containing A28330G region or oligonucleotide standards. (A) nPCR product amplified from TAB RNA sample (Sep 26, 2022); (B) short oligonucleotide at 1 nmol corresponding to variant mutation (GGG at 28328-28330 region); and (C) short oligonucleotide at 1 nmol corresponding to wildtype (GGA at 28328-28330 region). For each chromatograms, the first peak is single stranded DNA and the second peak is double stranded DNA (Huber and Oberacher 2001, Murray 1996). Total area of the two peaks at the *m/z* corresponding to single strand DNA were calculated for each strand.
